# ACUTE SEIZURES AFTER SPONTANEOUS INTRACEREBRAL HEMORRHAGE IN YOUNG INDIVIDUALS: 11-YEAR TRENDS AND ASSOCIATION WITH MORTALITY

**DOI:** 10.1101/2024.03.15.24304381

**Authors:** Alain Lekoubou, Austin Cohrs, Mariana Dejuk, Jinpyo Hong, Souvik Sen, Leonardo Bonilha, Vernon M. Chinchilli

**Affiliations:** Department of Neurology, Milton S. Hershey Medical Center, Pennsylvania State University; Department of Public Health Sciences, Pennsylvania State University; College of Medicine, Penn State University, Hershey, PA; University of South Carolina, Department of Neurology

**Keywords:** stroke, seizures, young, trends

## Abstract

**OBJECTIVE:** To describe trends in acute seizures (AS) among young individuals with spontaneous intracerebral hemorrhage (sICH) and association with mortality.

**BACKGROUND:** Acute seizures are frequent complications of sICH. The rate of sICH is rising among young Americans (18 to 60 years). Trends in AS incidence in this age group is largely unknown. Further, the association of AS with mortality has not been reported among young Americans.

**DESIGN/METHODS:** The Merative MarketScan® Commercial Claims and Encounters database, for the years 2005 through 2015, served as the data source for this study. This period was chosen as spontaneous ICH incidence increased among young individuals between 2005 and 2015. Our study population included patients aged 18 to 64 years with ICH identified using the International Classification of Diseases, Ninth and Tenth Revision (ICD-9/10) codes 430, 431, 432.0, 432.1, 432.9, I61, I61.0, I61.1, I61.2, I61.3, I61.4, I61.5, I61.6, I61.8, and I61.9, excluding those with a prior diagnosis of seizures (ICD-9/10 codes 345.x,780.3x, G40, G41, and R56.8). We computed yearly AS incidence, mortality (in patients with and without seizures), and analyzed trends. We applied a logistic regression model to determine the independent association of AS with mortality accounting for demographic and clinical variables.

**RESULTS:** Of 81,878 sICH patients, 7,611 (9.3%) developed AS. AS incidence increased linearly between 2005 (incidence rate: 8.1%) and 2015 (incidence rate: 11.0%), which represents a 26% relative increase (P for trends <0.0001. In-hospital mortality rate was 14.3% among those who developed AS and 11.5% among those who did not. Between 2005 and 2015, overall, in-hospital mortality decreased from 13.0% to 9.7% among patients without AS but remained unchanged among those with AS. Patients who developed AS were 10% more likely to die than those who did not (OR: 1.10, 95% confidence interval: 1.02-1.18).

**CONCLUSIONS:** Between 2005 and 2015, AS incidence increased by nearly 26% among young Americans with sICH. In-patient mortality remained unchanged among those who developed seizures but declined among those who did not. The occurrence of AS was independently associated with a 10% higher risk of in-hospital death. Future studies will test the benefit of treating AS to reduce mortality after sICH.

## Introduction

Spontaneous intracerebral hemorrhage (sICH), once considered primarily a disease of the elderly(1), is now increasingly afflicting young individuals across the United States, heralding a critical shift in stroke epidemiology. Each year, approximately 120,000 intracerebral hemorrhages occur in the United States(1). The incidence trends suggest that an increasing number of young individuals now develop spontaneous intracerebral hemorrhage (1, 2). Overall, the incidence of intracerebral hemorrhage has increased, and specifically between 2004 and 2018, an 11% increase in the incidence of intracerebral hemorrhage was observed. These incidence trends were largely driven by rising incidence among young and middle-aged Americans. During that period, the incidence of intracerebral hemorrhage increased significantly faster among those aged 18 to 44 years and 45 to 64 years compared to those aged ≥75 years(2). One of the public health implications of these findings is that acute sICH complications are likely to rise in this segment of the population.

Seizures are frequent early complications of stroke, occurring in up to 40% of patients after sICH when continuous electroencephalogram (EEG) monitoring is used(3, 4). Seizures are also more likely to develop in young adults than old individuals(5). The onset of seizures after ICH is associated with poor functional outcomes and increased risk of death(6, 7). The onset of seizures within the first days of ICH onset is also associated with a greater risk of subsequently developing epilepsy (recurrent spontaneous seizures)(8), which in turn carries an increased risk of long-term functional outcomes and cognitive deficits. The risk of dementia for example is multiplied by 2.5 when seizures develop any time after intracerebral(9).

To develop strategies to improve clinical outcomes among ICH patients with seizures in this segment of the population, it is important to have nationally representative estimates of differences in incidence across time. We are not aware of studies summarizing nationwide changes in the incidence of acute seizures after intracerebral hemorrhage in young individuals. Data on mortality trends and the association of seizures with death are also scant. In this analysis, we sought to provide nationally representative estimates of differences in seizures and mortality by seizure status between 2005 and 2015, a period during which a disproportionately higher increased incidence of ICH was observed in young individuals. The current analysis also aimed to quantify the impact of seizures on mortality after intracerebral hemorrhage in young individuals.

## Methods

### Standard protocol approvals, registrations, and patient consents

The study protocol was submitted to the Pennsylvania State College of Medicine institutional review board and was not considered to be human subjects research. All records contained within the database are fully de-identified. Thus, informed consent was waived.

### Data source, Study Design, and Case Identification

This was a retrospective cohort analysis spanning from January 2005 through December 2015. The Merative MarketScan® Commercial Claims and Encounters database was used as a data source. This database gathers reimbursed healthcare claims that include individual-level and longitudinal data from patients aged 0 to 64 years who are covered by private insurance plans across all 50 US states and the District of Columbia. The database contains inpatient and outpatient medical claims, enrollment information of enrollees, and prescription drug claims for approximately 540 million privately insured patients annually. Detailed information about the database is described elsewhere(10). Our study population included patients aged 18 years to 64 years hospitalized with a primary diagnosis of spontaneous intracerebral hemorrhage (s-ICH) identified using the International Classification of Diseases, Ninth and Tenth Revision (ICD-9/10) codes 430, 431, 432.0, 432.1, 432.9, I61, I61.0, I61.1, I61.2, I61.3, I61.4, I61.5, I61.6, I61.8, and I61.9, excluding those with a prior diagnosis of seizures (ICD-9/10 codes 345.x,780.3x, G40, G41, and R56.8) between January 2005 and December 2015.

### Statistical analyses

We used SAS statistical software version 9.4 (SAS Institute, Cary, NC, USA) for analyses. We computed the overall incidence of AS and compared patients who developed seizures with those who did not develop seizures using chi-square (χ^2^) and t-tests respectively for categorical and continuous variables. Patients with AS and those without AS were compared using the following variables: Age (in years), sex (male vs. female), geographic regions-1 (Northeast, North Central, South West, Unknown), geographic regions-2 (rural vs. urban), medical comorbidities (hypertension, diabetes, myocardial infarction, heart failure, atrial fibrillation, alcohol use, depression, coma), and surgical treatment (Craniotomy). The clinical variables were identified using ICD-9/10 codes available in the supplemental files. We compared yearly AS incidence change (P-value for trends). Since changes in AS incidence could be affected by an increased rate of detection using continuous EEG monitoring, we computed AS incidence trends separately in patients who had continuous EEG monitoring and those who did not have continuous EEG monitoring. Finally, to assess the independent association of acute seizures with inpatient mortality, we applied a logistic regression model accounting for demographic (age, sex, geographic regions) and clinical variables (history of alcohol use, history of diabetes, history of hypertension, history of myocardial infarction, history of congestive heart failure, history of atrial fibrillation, depression, presence of coma, and whether or not a craniotomy was performed). Statistical significance was set at <0.05.

### Data availability

All data used for this research are publicly available through IBM Watson Health MarketScan® Commercial Claims and Encounters database.

## Results

### General characteristics

Our study population comprised 81,878 patients with spontaneous intracerebral hemorrhage and no prior history of seizures. Out of these patients, 7,611 (9.3%) had seizures during hospitalization (Table 1). The mean number of days to seizure was 3.9 days and 75% of seizures occur within the first day of stroke (Figure 1). There were no differences in acute seizure distribution based on sex. AS was more frequent in the Northeast (11.9%) and less frequent in the West (8.0%), p-value <0.0001. Similarly, AS was more frequent in urban than in rural areas (9.4% vs.8.5%, p-value 0.0007). In general, except for a history of alcohol use, medical comorbidities were more frequent in patients with acute seizures, all p-value ≤0.0001. AS incidence was higher in patients in coma than those who were not in coma (19.1% vs. 7.9%, p-value <0.0001). Similarly, having craniotomy was associated with a higher incidence of seizures (12.9% vs. 9.2%, p-value <0.0001). AS incidence was also higher among patients who were on continuous EEG monitoring compared with those who were not (42.7% vs. 4.2%, p-value <0.0001). Finally, AS incidence was higher among patients who died in the hospital than those who survived (11.3% vs. 9.0%, p-value <0.0001) (Table 1).

**Table 1:** General characteristics of young patients with spontaneous intracerebral hemorrhage, comparing those with and those without acute seizures. MarketScan Database 2005-2015.

|  | No Seizure<br>(N=74267) | Seizure<br>(N=7611) | P-Value |
| --- | --- | --- | --- |
| Mean Age ± SD | 50.4 ± 11.6 | 49.4 ± 12.1 | <.0001 |
| Sex |  |  | 0.0806 |
| Male | 39,820 | 4001 |  |
| Female | 34,447 | 3610 |  |
| Geographic region 1 |  |  | <.0001 |
| Northeast | 11,637 | 1,569 |  |
| North Central | 17,988 | 1788 |  |
| South | 31,318 | 3,068 |  |
| West | 12,319 | 1069 |  |
| Unknown | 1,005 | 117 |  |
| Geographic region 2 |  |  | 0.0007 |
| Rural | 11,050 | 1022 |  |
| Urban | 63,217 | 6589 |  |
| Comorbidities |  |  |  |
| Diabetes | 12320 | 1237 | 0.4526 |
| Hypertension | 30800 | 3168 | 0.7977 |
| Congestive Heart Failure | 5939 | 858 | <.0001 |
| Myocardial Infarction | 2328 | 337 | <.0001 |
| Atrial Fibrillation | 4250 | 523 | <.0001 |
| Alcohol Use | 3681 | 378 | 0.9693 |
| Depression | 2678 | 341 | 0.0001 |
| Craniotomy |  |  |  |
| Yes | 1810 | 267 | <.0001 |
| Coma |  |  |  |
| Yes | 7968 | 1886 | <.0001 |
| EEG (CPT 95951) |  |  |  |
| Yes | 6178 | 4612 | <.0001 |
| Discharge Status |  |  |  |
| Died | 8564 | 1090 | <.0001 |

**Table 2:** Logistic regression analysis for the association of acute seizures with inpatient mortality among young patients with intracerebral hemorrhage. MarketScan database: 2005-2015.

|  | **Odds ratio** | **95% Confidence intervals** |  |
| --- | --- | --- | --- |
| **Acute seizures vs. No seizures** | 1.097 | 1.022 | 1.177 |
| **Age** | 1.021 | 1.019 | 1.023 |
| **SEX Male vs. Female** | 1.043 | 0.998 | 1.09 |
| **North Central vs. Northeast** | 1.074 | 1.001 | 1.153 |
| **South vs. Northeast** | 1.159 | 1.086 | 1.236 |
| **West vs. Northeast** | 1.056 | 0.977 | 1.141 |
| **Unknown vs. Northeast** | 0.921 | 0.752 | 1.128 |
| **Rural vs. Urban** | 1.126 | 1.061 | 1.195 |
| **History of diabetes mellitus** | 0.955 | 0.901 | 1.013 |
| **History of hypertension** | 0.706 | 0.674 | 0.739 |
| **History of congestive heart failure** | 1.299 | 1.209 | 1.395 |
| **History of acute myocardial infarction** | 2.053 | 1.866 | 2.258 |
| **History of atrial fibrillation** | 1.266 | 1.166 | 1.374 |
| **History of alcohol use** | 0.586 | 0.518 | 0.663 |
| **History of depression** | 0.494 | 0.426 | 0.573 |
| **Craniotomy** | 1.406 | 1.244 | 1.59 |
| **Coma** | 2.626 | 2.487 | 2.772 |

**Figure 1:**
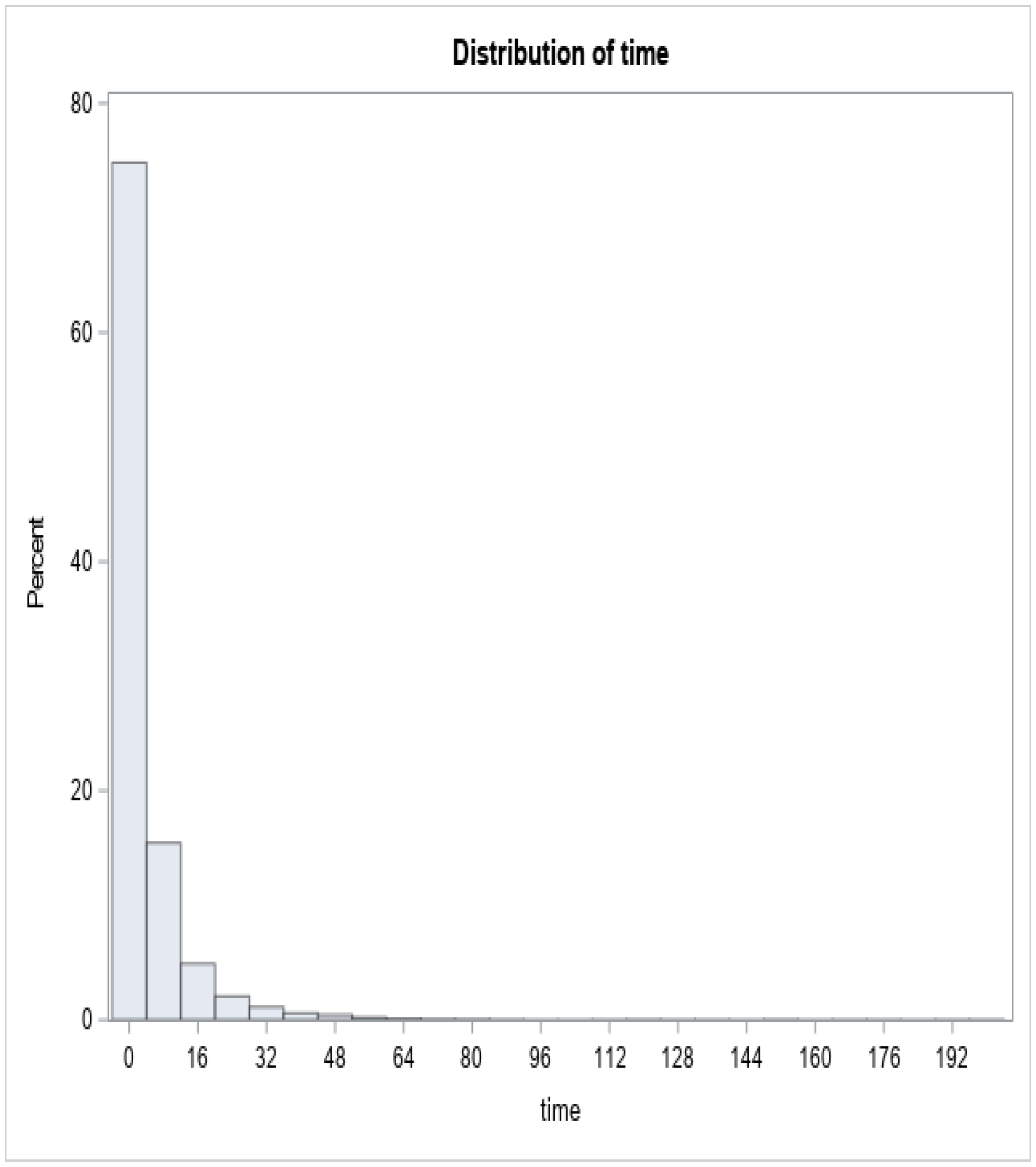
Distribution of time (days) from stroke to seizure.

### Trends analyses

Overall, seizure incidence increased from 8.1% in 2005 to 11% in 2015 (P-trends <0.001).

To test the hypothesis that the increase in seizure rate was not due to an increased detection rate, we computed seizure incidence trends separately in patients who had continuous EEG monitoring (1,608 patients) and those who did not have EEG (80,278). Overall, seizure detection did not statistically change between 2005 and 2015 (P for trends = 0.624). Among those who did not have EEG, AS incidence was 7.9% and increased to 8.5% in 2015 (P for trends = 0.0129) (Figure 2). In-hospital mortality decreased from 13.0% to 9.7% among patients who did not have AS (P for trends <0.0001), but remained unchanged among patients with AS (P for trends = 0.8045) (Figure 3).

**Figure 2:**
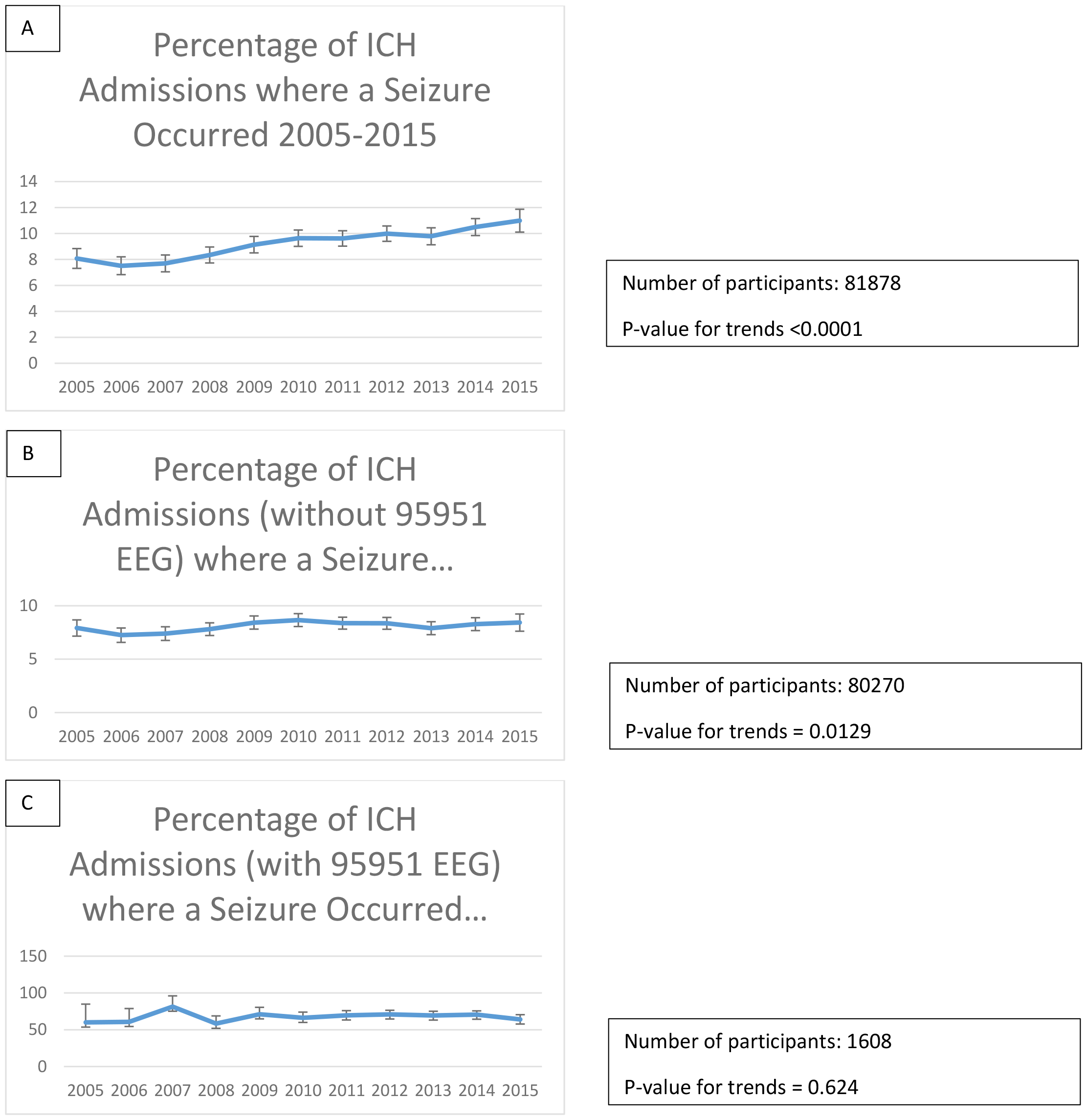
Trends in acute seizures rate among young patients with intracerebral hemorrhage. Marketscan databse: 2005-2015 A: All patients. B: Patients who did not have a continuous electroencephalogram (clinical diagnosis only) C: Patients who had a continuous electroencephalogram study.

**Figure 3:**
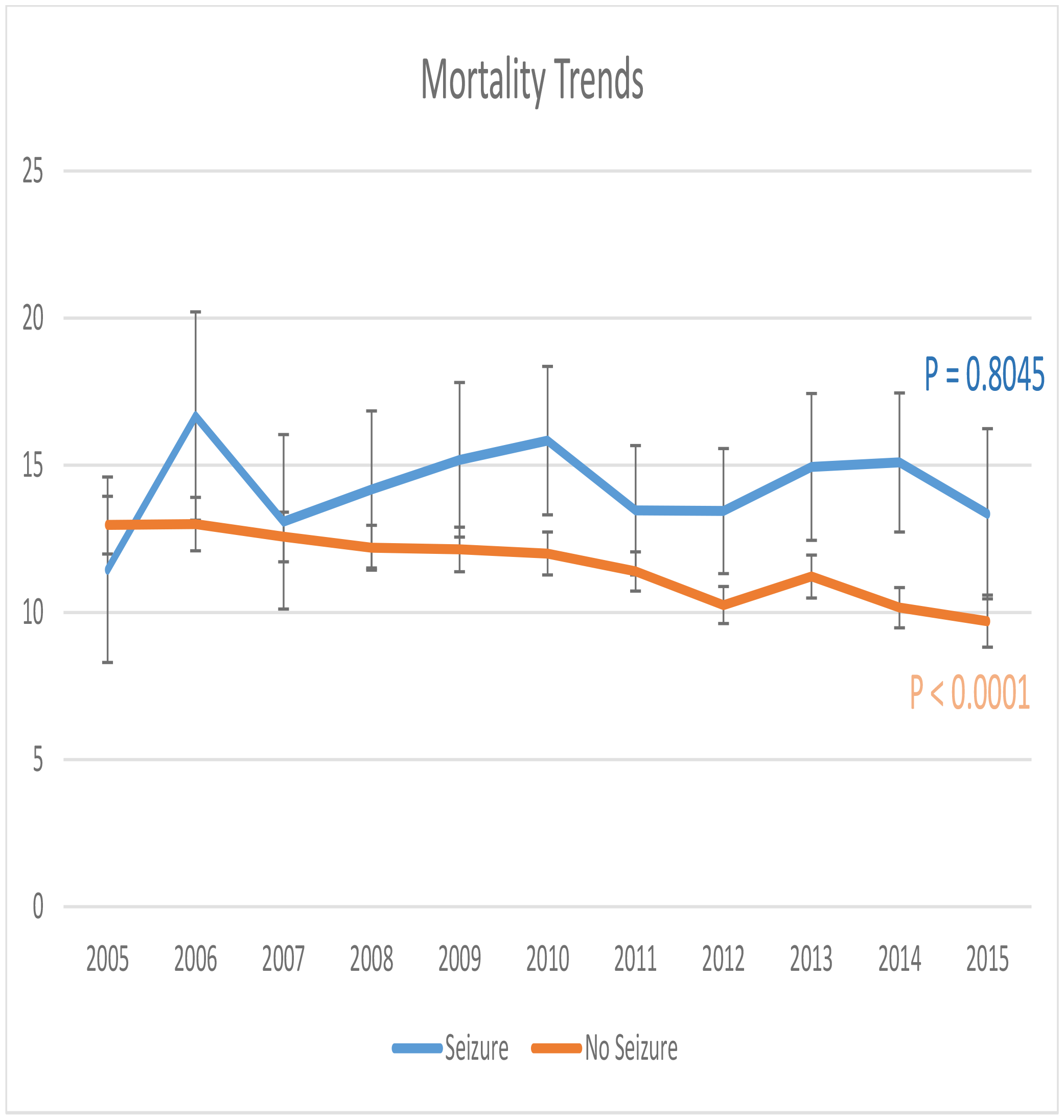
Mortality trends in young patients (18-64 years) with intracerebral hemorrhage. Marketscan database: 2005-2015

### Association of acute seizures with mortality

A total of 9,654 in-hospital deaths were recorded between 2005 and 2015 among young patients hospitalized for intracerebral hemorrhage. Of these deaths, 1,090 (14.3%) occurred in those who had AS and 8,564 (11.50%) among those who did not have AS, P <0.0001%. After adjusting for sociodemographic factors/hospital-related (age, sex, geographic location), medical comorbidities, craniotomy/coma, acute seizures remained associated with a 10% increased odds of in-hospital death (OR: 1.10, 95% confidence interval: 1.02-1.18) (Figure 2).

## Discussion

The current analysis shows a 26% increase in seizure incidence among young patients hospitalized for sICH between 2005 and 2015. During the same period, in-hospital mortality did not change among young sICH patients with seizures but decreased among those who did not develop seizures. We report a 10% independent increased risk of in-hospital death associated with seizures in young patients with sICH.

Seizures are frequent complications of sICH with suggestions from large epidemiologic hospital-based cohorts that it is more prevalent in young than older individuals(5). The observed increase in seizure incidence between 2005 and 2015 highlights the substantial public health implications of the reported rise in sICH incidence among this demographic.” Detecting and treating seizures after intracerebral hemorrhage could potentially reduce long-term complications of sICH, as seizures are associated with significant long-term complications such as cognitive impairment. For example, in another paper by our team, we showed that seizures occurring any time after sICH were associated with a 2.5-fold increased risk of developing dementia(9). Acute seizures after sICH have also been associated in some studies with a poor outcome(3, 6, 11). In line with these previous reports, for the first time, we have shown that in this specific segment of the population, i.e. young patients with intracerebral hemorrhage, seizures were associated with a 10% increased risk of death after controlling for factors such as stroke severity (coma) and craniotomy which are known to be strongly associated with mortality after sICH. Our study also points out the need for early seizure detection in this population. One in three seizures occurred within 24 hours of stroke onset. Further, we showed that in patients who had continuous EEG, the seizure rate was 42%, a figure similar to acute seizure incidence reported in studies that used continuous EEG for seizure detection(3, 4). The clinical implication of these findings is that patients with sICH should be closely monitored within the first hours of stroke for seizure detection and treatment. It can also be argued that clinicians should have a low threshold for using continuous EEG monitoring. Prediction models that will help identify patients with the highest risk of developing acute seizures and benefit from antiseizure medications without suffering the deleterious effects of these drugs are highly needed. These tools will help rationalize resource utilization such as continuous EEG monitoring and EEG/epilepsy staff. Independent predictors of seizures after intracerebral hemorrhage have been identified and include young age, large hemorrhage, cortical hemorrhage, and a prior history of intracerebral hemorrhage(12). Although these individual risk factors are known, they do not inform the individual risk of developing early seizures following intracerebral hemorrhage. We are not aware of any models that included patients with subclinical seizures (diagnosed with continuous EEG monitoring) and accounted for the time from stroke onset to seizures.

## Limitations

Our study has potential limitations. Although Marketscan covers all 50 states and the District of Columbia, it is possible that we did not account for all hospitalizations of young patients with intracerebral hemorrhage as the database contain only patients with commercial insurance. Young patients are however more likely to have commercial insurance than elderly patients many of whom are on MEDICARE. The diagnoses of intracerebral hemorrhage and seizures were based on ICD codes. Stroke diagnoses based on ICD-codes have been used in epidemiologic study with accuracy as high as 100%(2). Seizures diagnosis using ICD codes also has a good sensitivity and specificity(13, 14). Our estimates are close to those provided in studies that relied on clinical diagnoses. Finally, Marketscan does not contain data on race and ethnicity, hence racial/ethnic differences could not be assessed. Despite these limitations, our study is the largest on young patients with sICH evaluating changes in seizure incidence and related mortality covering a period of more than 10 years.

## Conclusion

AS incidence has increased between 2005 and 2015 among young patients hospitalized for sICH, a period of time during which ICH also increased in this age group. In-hospital mortality did not change among patients those who had seizures unlike patients who did not have seizure. Developing AS was associated with a 10% increase in the risk of in-hospital death. There is an urgent need to develop national strategies for early identification and treatment of seizures in the acute phase of sICH that will account for the particular vulnerability of young individuals.

## References

1. Tsao CW, Aday AW, Almarzooq ZI, Anderson CAM, Arora P, Avery CL, et al. Heart Disease and Stroke Statistics-2023 Update: A Report From the American Heart Association. Circulation. 2023;147(8):e93–e621.

2. Bako AT, Pan A, Potter T, Tannous J, Johnson C, Baig E, et al. Contemporary Trends in the Nationwide Incidence of Primary Intracerebral Hemorrhage. Stroke. 2022;53(3):e70–e4.

3. Claassen J, Jette N, Chum F, Green R, Schmidt M, Choi H, et al. Electrographic seizures and periodic discharges after intracerebral hemorrhage. Neurology. 2007;69(13):1356–65.

4. Peter-Derex L, Philippeau F, Garnier P, Andre-Obadia N, Boulogne S, Catenoix H, et al. Safety and efficacy of prophylactic levetiracetam for prevention of epileptic seizures in the acute phase of intracerebral haemorrhage (PEACH): a randomised, double-blind, placebo-controlled, phase 3 trial. Lancet Neurol. 2022;21(9):781–91.

5. Merkler AE, Gialdini G, Lerario MP, Parikh NS, Morris NA, Kummer B, et al. Population-Based Assessment of the Long-Term Risk of Seizures in Survivors of Stroke. Stroke. 2018;49(6):1319–24.

6. Law ZK, England TJ, Mistri AK, Woodhouse LJ, Cala L, Dineen R, et al. Incidence and predictors of early seizures in intracerebral haemorrhage and the effect of tranexamic acid. Eur Stroke J. 2020;5(2):123–9.

7. Vespa PM, O’Phelan K, Shah M, Mirabelli J, Starkman S, Kidwell C, et al. Acute seizures after intracerebral hemorrhage: a factor in progressive midline shift and outcome. Neurology. 2003;60(9):1441–6.

8. Vergara Lopez S, Ramos-Jimenez C, Adrian de la Cruz Reyes L, Kevin Galindo Ruiz A, Armando Baigts Arriola L, Manuel Moncayo Olivares J, et al. Epilepsy diagnosis based on one unprovoked seizure and >/=60% risk. A systematic review of the etiologies. Epilepsy Behav. 2021;125:108392.

9. Lekoubou A, Ba DM, Nguyen C, Liu G, Leslie DL, Bonilha L, et al. Poststroke Seizures and the Risk of Dementia Among Young Stroke Survivors. Neurology. 2022;99(4):e385–e92.

10. Truven Health Analytics potIWHb. The Truven Health MarketScan databases for health services researchers. [Available from: Available at: https://www.ibm.com/downloads/cas/6KNYVVQ2..

11. Naidech AM, Weaver B, Maas M, Bleck TP, VanHaerents S, Schuele SU. Early Seizures Are Predictive of Worse Health-Related Quality of Life at Follow-Up After Intracerebral Hemorrhage. Crit Care Med. 2021;49(6):e578–e84.

12. De Herdt V, Dumont F, Henon H, Derambure P, Vonck K, Leys D, et al. Early seizures in intracerebral hemorrhage: incidence, associated factors, and outcome. Neurology. 2011;77(20):1794–800.

13. Jette N, Beghi E, Hesdorffer D, Moshe SL, Zuberi SM, Medina MT, et al. ICD coding for epilepsy: past, present, and future--a report by the International League Against Epilepsy Task Force on ICD codes in epilepsy. Epilepsia. 2015;56(3):348–55.

14. Jette N, Reid AY, Quan H, Hill MD, Wiebe S. How accurate is ICD coding for epilepsy? Epilepsia. 2010;51(1):62–9.

